# A Large-Scale Observational Study on the Temporal Trends and Risk Factors of Opioid Overdose: Real-World Evidence for Better Opioids

**DOI:** 10.1101/2020.10.08.20208678

**Authors:** Jianyuan Deng, Wei Hou, Xinyu Dong, Janos Hajagos, Mary Saltz, Joel Saltz, Fusheng Wang

## Abstract

**Background:** The United States is in the midst of an opioid overdose epidemic. We evaluated the temporal trends and risk factors of inpatient opioid overdose. Based on the opioid overdose patterns, we further examined the innate properties underlying less overdose events.

**Methods:** We conducted a retrospective cross-sectional study based a large-scale inpatient electronic health records database, Cerner Health Facts^®^. We included patients admitted between January 1, 2009 and December 31, 2017. Opioid overdose prevalence by year, demographics and prescription opioid exposures.

**Results:** A total of 4,720,041 patients with 7,339,480 inpatient encounters were retrieved from Cerner Health Facts^®^. Among them, 30.2% patients were aged 65+, 57.0% female, 70.1% Caucasian, 42.3% single, 32.0% from South and 80.8% in urban area. From 2009 to 2017, annual opioid overdose prevalence per 1,000 patients significantly increased from 3.7 to 11.9 with an adjusted odds ratio (aOR): 1.16, 95% confidence interval (CI): [1.15-1.16]. Comparing to the major demographic counterparts above, being in 1) age group: 41-50 (overall aOR 1.36, 95% CI: [1.31-1.40]) or 51-64 (overall aOR 1.35, 95% CI: [1.32-1.39]), marital status: divorced (overall aOR 1.19, 95% CI: [1.15-1.23]), 3) census region: West (overall aOR 1.32, 95% CI: [1.28-1.36]), were significantly associated with higher odds of opioid overdose. Prescription opioid exposures were also associated with increased odds of opioid overdose, such as meperidine (overall aOR 1.09, 95% CI: [1.06-1.13]) and tramadol (overall aOR 2.20. 95% CI: [2.14-2.27]). Examination on the relationships between opioid agonists’ properties and their association strengths, aORs, in opioid overdose showed that lower aORs values were significantly associated with 1) high molecular weight, 2) negative interaction with multi-drug resistance protein 1 (MDR1) or positive interaction with cytochrome P450 3A4 (CYP3A4) and 3) negative interaction with delta opioid receptor (DOR) or kappa opioid receptor (KOR).

**Conclusions:** The significant increasing trends of opioid overdose at the inpatient care setting from 2009 to 2017 indicated an ongoing need of efforts to combat the opioid overdose epidemic in the US. Risk factors associated with opioid overdose included patient demographics and prescription opioid exposures. Different prescription opioids were associated with opioid overdose to different extents, indicating a necessity to better differentiate them during prescribing practice. Moreover, there are physicochemical, pharmacokinetic and pharmacodynamic properties underlying less overdose events, which can be utilized to develop better opioids.

**Key Points:**

- There were significant increasing trends of opioid overdose at the US inpatient care setting from 2009 to 2017, showing an ongoing need for opioid overdose prevention.
- Different prescription opioids were associated with opioid overdose to different extents, indicating a necessity to differentiate prescription opioids during prescribing.
- The optimal properties underlying less overdose events mined from the large-scale, real-world electronic health records hold high potential to guide the development of better opioids with reduced overdose effects.

## 1 Introduction

Over the past two decades, drug overdose has been a leading cause of injury-related deaths in the United States (US), of which 70% involved illicit or prescription opioids [1]. Prescription opioids are among the most effective drugs to treat pain, which are ligands to the endogenous mu opioid receptor (MOR) and can exert agonistic, partially agonistic or antagonistic effect [2]. When activated by agonists, MOR can mediate analgesic effects as well as modulate respiratory responses [3]. However, lethal respiratory depression can happen in case of overdose [4]. In 2018, opioid overdose was attributed to 47,761 deaths, which imposed an enormous public health burden on the US [5, 6]. The epidemic of opioid overdose is dynamic and complex [7, 8]. As opined by Jalal et al, the current epidemic can be a recent manifestation of an ongoing longer-term process [8]. Close monitoring the temporal trends of opioid overdose is, therefore, crucial for developing and evaluating relevant policies to prevent and control the epidemic [8-12]. Furthermore, despite the complexity of the opioid overdose epidemic, there exist some patterns on patient demographics and opioid prescriptions, which can provide essential knowledge for the planning of effective prevention measures [12-15].

By now, various guidelines and programs have already been launched to curb opioid prescribing [16-18]. Notwithstanding the lethal respiratory depression effect, opioids are still among the most commonly prescribed medications, presumably due to that pain is the one of most common reasons for patients to visit their doctors [19, 20]. To combat the overdose epidemic, better opioid agonists with reduced overdose effects are indisputably needed [21]. Novel treatments have been proposed based on optimizing their innate properties. For example, one selective MOR agonist was shown to be both effective for analgesia and devoid of respiratory depression [22]. Another MOR agonist was also reported to have low abuse potential due to a reduced rate of entry across the blood brain barrier (BBB) [23]. Nonetheless, whether these new chemical entities will eventually work in human subjects remains unclear yet [21]. In fact, new drug development is well known for its low success rate [24]. Thus, given the severity and urgency of the opioid overdose epidemic, early-stage optimization strategies for the lead compounds are the key to successfully bring better opioids into clinical use, which include physicochemical, pharmacokinetic and pharmacodynamic aspects [25].

Recently, real-world data (RWD), such as Electronic Health Records (EHRs), has received substantial attention for large-scale drug safety study [26]. Real-world evidence (RWE) generated from RWD holds high potential to guide drug discovery and subsequent drug development [27, 28]. In the case of opioid overdose, the fundamental question would be “What are the optimal properties underlying opioids with less overdose effects?”. The properties associated with less overdose events in a large human population can serve as the valid target product profiles to clearly define the desired attributes during early drug discovery, thereby maximizing the success rate of better opioid agonists eventually getting approved [29].

Hence, in this large-scale observational study on opioid overdose, we firstly examined the temporal trends of opioid overdose and its risk factors, including patient demographics and prescription opioid exposures based on a large-scale EHRs database. Secondly, we examined opioid agonists’ relevant properties underlying lower association strengths in opioid overdose so as to generate RWEs on optimal properties for better opioids with less overdose events.

## 2 Methods

### 2.1 Data and Measurements

This retrospective cross-sectional study was based on Cerner Health Facts^®^, one of the largest EHRs databases in the United States. Health Facts^®^ stores real-world, de-identified patient data, such as records on encounters, diagnoses and medications, from over 600 healthcare facilities. A recent study showed that Health Facts^®^ and the Healthcare Cost and Utilization Project Nationwide Inpatient Sample have similar distribution across all data elements [30].

For the assembly of our study dataset, we retrieved patients at the inpatient care setting admitted between January 1, 2009 and December 31, 2017. To identify patients with opioid overdose, we used the International Classification of Diseases codes, Ninth Revision (ICD-9) and Tenth Revision (ICD-10), for poisoning or adverse effect by opium, heroin, methadone, other related narcotics, etc (Electronic Supplementary Material, Table S1) [31]. Patients with at least one relevant diagnosis code in all inpatient encounters within a year were added to the opioid overdose cohort in that year. Other independent variables are described as follows.

### 2.2 Patient Demographics

We measured patient demographics, including age group (<18, 18-30, 31-40, 41-50, 51-64, 65+), gender (female, male), race/ethnicity (African American, Asian, Caucasian, Hispanic, Native American, others), marital status (divorced, married, single, widowed, others), census region (Midwest, Northeast, South, West) and urbanicity (rural, urban). For patients with varied demographics, such as migration between rural and urban areas, records from their first encounter were used.

### 2.3 Prescription Opioids

For prescription opioids, we categorized them by their action types on MOR [2], namely, 1) opioid agonists: codeine, fentanyl and morphine, among others, 2) opioid partial agonists: buprenorphine, butorphanol and nalbuphine, and 3) opioid antagonist: naloxone. For each category, we collected the corresponding FDA-approved active ingredients from DrugBank 5.1.6, which were then mapped to medications containing the active ingredients in Health Facts^®^ [32]. Illicit opioids and opioids withdrawn from market were excluded, such as diamorphine and propoxyphene. To ensure sufficient statistical significance, prescription opioids with annual prevalence less than 1‰ were also excluded from analyses, such as dihydrocodeine, naltrexone and pentazocine. Medications in all inpatient encounters within one year were aggregated and a patient was defined as exposed if medications containing the active ingredient were prescribed in that year, otherwise as non-exposed.

For prescription opioids’ innate properties, they were also collected from DrugBank 5.1.6 [32], including, 1) physicochemical properties underlying BBB permeability: lipophilicity (logP), polar surface area (PSA) and molecular weight (MW) [33], 2) pharmacokinetic interaction with efflux transporters and metabolizing enzymes: multi-drug resistance protein 1 (MDR1), cytochromes P450 2D6 (CYP2D6) and 3A4 (CYP3A4) [34-36], and 3) pharmacodynamic interaction with receptors: delta opioid receptor (DOR), kappa opioid receptor (KOR) and N-methyl-D-aspartate receptor (NMDAR) [22, 37].

### 2.2 Statistical Analyses

Demographic characteristics were determined using descriptive analyses. Proportions of opioid overdose, i.e., opioid overdose prevalence, among all patients and patient subgroups divided by demographics, were calculated on a yearly basis.

Inferential analyses were conducted to examine the temporal trends of opioid overdose and risk factors by calculating the adjusted odds ratios (aORs) and 95% confidence intervals (CIs). Firstly, for each patient subgroup stratified by demographics, multivariable logistic regression was used with 1) the inclusion of whether a patient was diagnosed as opioid overdose within a year as the dependent variable, and 2) year and a demographic factor as independent variables plus an interaction item between the two independent variables to examine the temporal trends, further adjusted for all demographics (e.g., age_group, gender, race/ethnicity, marital status, census region and rural/urban area). Secondly, for risk factors, including demographics and prescription opioid exposures, multivariable logistic regression adjusted for all demographics was also performed based on both data in each year for the yearly aORs and data across all years for the overall aORs. For demographics, age group in 65+, gender as female, race/ethnicity as Caucasian, marital status as single, census region in south and urban area were set as the reference. For prescription opioid exposures, the non-exposed was set as the reference.

To examine the relationship between MOR agonists’ innate properties and their aORs (i.e., association strengths) in opioid overdose, power analysis was conducted to only exploit aORs with statistical power greater than 80% [38, 39]. For continuous physicochemical properties, we first discretized them into two groups, low or high, with median as the cutoff. Then, Wilcoxon test was used to examine the relationship between aORs of opioid agonists and their attributes in the physicochemical, pharmacokinetic and pharmacodynamic aspects. Dataset was processed in Python 3.7.3. Statistical analyses were conducted in R 3.6.1, packages: “stats”, “epiR”. All data analyses were performed between Feb 1, 2020 and Sept 1, 2020.

## 3 Results

### 3.1 Patient Characteristics

Between 2009 and 2017, a total of 7,339,480 inpatient encounters for 4,720,041 patients were retrieved from Health Facts^®^. Among them, 1,423,744 (30.2%) patients were in the 65+ age group, 2,689,898 (57.0%) patients were female, 3,310,471 (70.1%) patients were Caucasian, 1,995,380 patients (42.3%) were single, 1,511,746 patients (32.0%) were from South and 3,813,817 patients (80.8%) were in urban area. Encounter counts for patients were summarized in Electronic Supplementary Material, Figure S1.

### 3.2 Temporal Trends of Opioid Overdose

Annual opioid overdose prevalence from 2009 to 2017 stratified by demographics was detailed in Table 1. There were significant increasing trends of opioid overdose. From 2009 to 2017, the overall opioid overdose prevalence increased from 3.7‰ to 11.9‰ (3.2-fold increase; aOR: 1.16, 95% CI: [1.15-1.16]; p < 0.0001). The increasing trends in all patient subgroups were also significant. Among them, patient subgroup from South had a 6.3-fold increase (from 2.0‰ to 12.9‰; aOR: 1.22, 95% CI: [1.20-1.22]; p < 0.0001), followed by Asian patients (5.2-fold increase: from 1.2‰ to 6.0‰; aOR: 1.20, 95% CI: [1.14-1.27]; p < 0.0001), African American patients (4.8-fold increase: from 2.2‰ to 10.8‰; aOR: 1.20, 95% CI: [1.19-1.22]; p < 0.0001) and patients in rural area (4.2-fold increase: from 2.1‰ to 8.8‰; aOR: 1.14, 95% CI [1.13-1.16]; p < 0.0001). These prominent trends indicated that prevention and control measures of opioid overdose at the US inpatient care setting had been be inadequate over the years.

**Table 1.** Temporal Trends of Opioid Overdose Stratified by Demographics from 2009 to 2017.

| Year | No. of Opioid Overdose Patients (Opioid Overdose Prevalence, ‰) |  |  |  |  |  |  |  |  | aORs |
| --- | --- | --- | --- | --- | --- | --- | --- | --- | --- | --- |
|  | 2009 | 2010 | 2011 | 2012 | 2013 | 2014 | 2015 | 2016 | 2017 | [95% CI] |
| **Total** | 395,040 | 298,317 | 529,611 | 645,504 | 530,520 | 1,040,515 | 1,034,306 | 771,903 | 569,153 |  |
| **All Patients** | 1,477<br>(3.7) | 1,240<br>(4.2) | 2,749<br>(5.2) | 4,022<br>(6.2) | 3,844<br>(7.2) | 7,689<br>(7.4) | 8,313<br>(8.0) | 8,527<br>(11.0) | 6,783<br>(11.9) | 1.16 ****<br>[1.15-1.16] |
| **Age Groups** |  |  |  |  |  |  |  |  |  |  |
| <18 | 86<br>(1.2) | 77<br>(1.4) | 77<br>(0.9) | 162<br>(1.5) | 102<br>(1.5) | 307<br>(1.6) | 305<br>(1.6) | 200<br>(1.7) | 169<br>(2.3) | 1.08 ****<br>[1.06-1.10] |
| 18-30 | 207<br>(4.2) | 165<br>(4.5) | 366<br>(5.4) | 448<br>(5.1) | 406<br>(5.5) | 790<br>(5.6) | 900<br>(6.5) | 793<br>(8.0) | 612<br>(9.2) | 1.10 ****<br>[1.09-1.12] |
| 31-40 | 147<br>(4.2) | 124<br>(4.8) | 291<br>(5.7) | 387<br>(6.1) | 363<br>(6.8) | 714<br>(7.0) | 804<br>(7.8) | 774<br>(9.9) | 583<br>(10.3) | 1.12 ****<br>[1.10-1.14] |
| 41-50 | 192<br>(5.0) | 212<br>(7.5) | 474<br>(8.7) | 613<br>(9.6) | 545<br>(10.8) | 1,016<br>(11.3) | 1,000<br>(11.4) | 1,063<br>(15.8) | 839<br>(16.9) | 1.14 ****<br>[1.13-1.15] |
| 51-64 | 357<br>(5.0) | 273<br>(5.0) | 700<br>(6.6) | 1,105<br>(8.9) | 1,102<br>(10.5) | 2,152<br>(11.2) | 2,339<br>(12.3) | 2,506<br>(16.5) | 2,036<br>(17.3) | 1.18 ****<br>[1.17-1.19] |
| 65+ | 488<br>(3.8) | 389<br>(4.0) | 841<br>(5.0) | 1,307<br>(6.6) | 1,326<br>(7.4) | 2,710<br>(8.3) | 2,965<br>(9.2) | 3,191<br>(12.4) | 2,544<br>(12.4) | 1.18 ****<br>[1.17-1.18] |
| **Gender** |  |  |  |  |  |  |  |  |  |  |
| Female | 841<br>(3.7) | 692<br>(4.0) | 1,560<br>(5.2) | 2,314<br>(6.2) | 2,235<br>(7.3) | 4,448<br>(7.4) | 4,771<br>(8.1) | 4,873<br>(11.2) | 3,836<br>(12.1) | 1.16 ****<br>[1.15-1.17] |
| Male | 636<br>(3.9) | 548<br>(4.3) | 1,187<br>(5.2) | 1,708<br>(6.2) | 1,609<br>(7.2) | 3,240<br>(7.3) | 3,542<br>(8.0) | 3,654<br>(11.0) | 2,947<br>(11.7) | 1.15 ****<br>[1.14-1.16] |
| **Race** |  |  |  |  |  |  |  |  |  |  |
| African American | 143 | 120 | 371 | 517 | 389 | 821 | 975 | 1,060 | 1,051 | 1.20 **** |
|  | (2.2) | (2.4) | (3.9) | (4.6) | (4.6) | (5.6) | (6.6) | (9.0) | (10.8) | [1.19-1.22] |
| Asian | 6 | 2 | 17 | 29 | 37 | 65 | 90 | 98 | 71 | 1.20 **** |
|  | (1.2) | (0.5) | (2.2) | (2.5) | (3.4) | (3.1) | (4.2) | (5.2) | (6.0) | [1.14-1.27] |
| Caucasian | 1,240 | 1,054 | 2,198 | 3,216 | 3,130 | 6,207 | 6,604 | 6,717 | 5,068 | 1.15 **** |
|  | (4.3) | (4.9) | (5.9) | (7.3) | (8.5) | (8.4) | (9.1) | (12.5) | (13.4) | [1.14-1.16] |
| Hispanic | 23 | 16 | 39 | 41 | 19 | 44 | 29 | 25 | 17 | 1.14 **** |
|  | (2.4) | (2.2) | (2.9) | (3.7) | (4.0) | (4.3) | (3.5) | (6.9) | (6.1) | [1.08-1.20] |
| Native American | 3 | 5 | 14 | 25 | 51 | 123 | 137 | 135 | 92 | 1.20 **** |
|  | (3.0) | (7.8) | (3.6) | (3.9) | (7.3) | (7.7) | (8.7) | (15.2) | (11.6) | [1.14-1.26] |
| Others | 62 | 43 | 110 | 194 | 218 | 429 | 478 | 492 | 484 | 1.14 **** |
|  | (2.4) | (2.3) | (2.8) | (3.1) | (4.1) | (4.1) | (4.2) | (5.6) | (6.7) | [1.11-1.16] |
| **Marital Status** |  |  |  |  |  |  |  |  |  |  |
| Divorced | 153 | 117 | 364 | 502 | 491 | 936 | 1,045 | 1,082 | 850 | 1.16 **** |
|  | (6.6) | (6.6) | (9.0) | (10.6) | (12.1) | (13.0) | (14.9) | (19.4) | (19.7) | [1.14-1.17] |
| Married | 425 | 339 | 980 | 1,443 | 1,400 | 2,733 | 2,975 | 3,247 | 2,587 | 1.18 **** |
|  | (3.3) | (3.6) | (5.1) | (6.1) | (6.8) | (7.2) | (8.1) | (11.4) | (12.1) | [1.17-1.18] |
| Single | 403 | 328 | 855 | 1,338 | 1,242 | 2,583 | 2,799 | 2,723 | 2,230 | 1.13 **** |
|  | (3.0) | (3.2) | (4.3) | (5.1) | (6.2) | (5.9) | (6.3) | (8.4) | (9.8) | [1.12-1.14] |
| Widowed | 152 | 87 | 390 | 535 | 562 | 1,030 | 1,059 | 1,078 | 766 | 1.16 **** |
|  | (3.7) | (2.9) | (6.2) | (7.3) | (8.8) | (9.4) | (10.2) | (13.4) | (12.4) | [1.15-1.18] |
| Others | 344 | 369 | 160 | 204 | 149 | 407 | 435 | 397 | 350 | 1.16 **** |
|  | (5.0) | (6.7) | (4.5) | (7.1) | (8.4) | (9.2) | (9.5) | (14.5) | (15.1) | [1.14-1.17] |
| **Census Region** |  |  |  |  |  |  |  |  |  |  |
| Midwest | 328 | 272 | 564 | 656 | 625 | 1,409 | 1,703 | 1,934 | 1,070 | 1.18 **** |
|  | (3.6) | (4.2) | (4.7) | (5.9) | (6.6) | (6.9) | (7.4) | (13.3) | (11.9) | [1.17-1.19] |
| Northeast | 828 | 736 | 961 | 1,412 | 835 | 2,155 | 1,720 | 1,210 | 1,143 | 1.11 **** |
|  | (4.7) | (5.3) | (5.7) | (6.5) | (8.2) | (7.9) | (7.8) | (9.2) | (10.9) | [1.10-1.12] |
| South | 201 | 185 | 902 | 987 | 1,020 | 1,990 | 2,519 | 3,109 | 3,296 | 1.22 **** |
|  | (2.0) | (2.4) | (4.9) | (5.3) | (5.6) | (6.2) | (7.9) | (11.7) | (12.9) | [1.20-1.22] |
| West | 110 | 47 | 322 | 967 | 1,364 | 2,135 | 2,371 | 2,274 | 1,274 | 1.11 **** |
|  | (4.2) | (2.6) | (5.7) | (7.3) | (8.9) | (8.8) | (8.9) | (9.9) | (10.7) | [1.10-1.12] |
| **Urbanicity** |  |  |  |  |  |  |  |  |  |  |
| Rural | 112 | 117 | 435 | 777 | 528 | 1,404 | 1,494 | 1,172 | 967 | 1.14 **** |
|  | (2.1) | (2.2) | (5.0) | (5.7) | (5.5) | (6.7) | (6.9) | (8.0) | (8.8) | [1.13-1.16] |
| Urban | 1,365 | 1,123 | 2,314 | 3,245 | 3,316 | 6,285 | 6,819 | 7,355 | 5,816 | 1.16 **** |
|  | (4.0) | (4.6) | (5.2) | (6.4) | (7.6) | (7.6) | (8.3) | (11.8) | (12.7) | [1.15-1.16] |

\* p < 0.05; \*\* p < 0.01; \*\*\* p < 0.001; \*\*\*\* p < 0.0001.

### 3.3 Demographics as Risk Factors for Opioid Overdose

Demographics as risk factors for opioid overdose were summarized in Table 2. Compared to patients aged 65+, patients aged 41-50 (overall aOR: 1.36, 95% CI [1.31-1.40]; p < 0.0001) or 51-64 (overall aOR: 1.35, 95% CI [1.32-1.39]; p < 0.0001) had higher odds of opioid overdose whereas patients aged <18 (overall aOR: 0.18, 95% CI [0.17-0.19]; p < 0.0001) or 18-30 (overall aOR: 0.74, 95% CI [0.71-0.76]; p < 0.0001) had much lower odds. When comparing to single patients, patients who were divorced (overall aOR: 1.19, 95% CI [1.15-1.23]; p < 0.0001) had higher odds of opioid overdose whereas the odds were lower for patients who were married (overall aOR: 0.73, 95% CI [0.71-0.75]; p < 0.0001) or widowed (overall aOR: 0.89, 95% CI [0.86-0.93]; p < 0.0001). For patients in rural area (overall aOR: 0.84, 95% CI [0.82-0.86]; p < 0.0001), their odds of opioid overdose were lower than patients in urban area. Note that the yearly aORs were also varying. For example, across all years, male patients (overall aOR: 0.98, 95% CI [0.96-1.00]; p = 0.05) had comparable odds with female patients. However, male patients were more likely to have opioid overdose in 2009 whereas they became comparably or less likely to have it in subsequent years. Similarly, patients from Midwest, Northeast or West census regions also exhibited varying aORs of opioid overdose from 2009 to 2017, which partially reflected the changing dynamics of the opioid overdose epidemic [8].

**Table 2.** Demographics as Risk Factors for Opioid Overdose and Adjusted Odds Ratios from 2009 to 2017.

| Year | Adjusted Odds Ratios [95% CI] |  |  |  |  |  |  |  |  |  |
| --- | --- | --- | --- | --- | --- | --- | --- | --- | --- | --- |
|  | 2009 | 2010 | 2011 | 2012 | 2013 | 2014 | 2015 | 2016 | 2017 | Overall |
| **Age Groups <sup>a</sup>** |  |  |  |  |  |  |  |  |  |  |
| <18 | 0.28 ****<br>[0.21-0.36] | 0.32 ****<br>[0.24-0.42] | 0.20 ****<br>[0.15-0.25] | 0.22 ****<br>[0.18-0.26] | 0.21 ****<br>[0.17-0.26] | 0.18 ****<br>[0.16-0.21] | 0.17 ****<br>[0.15-0.19] | 0.14 ****<br>[0.12-0.16] | 0.19 ****<br>[0.16-0.22] | 0.18 ****<br>[0.17-0.19] |
| 18-30 | 1.10<br>[0.91-1.32] | 1.07<br>[0.88-1.31] | 1.24 **<br>[1.08-1.43] | 0.81 ***<br>[0.72-0.91] | 0.78 ****<br>[0.69-0.88] | 0.67 ****<br>[0.61-0.73] | 0.72 ****<br>[0.66-0.78] | 0.66 ****<br>[0.60-0.72] | 0.74 ****<br>[0.67-0.82] | 0.74 ****<br>[0.71-0.76] |
| 31-40 | 1.19<br>[0.98-1.44] | 1.22<br>[0.99-1.51] | 1.37 ****<br>[1.18-1.58] | 1.01<br>[0.89-1.14] | 1.03<br>[0.91-1.16] | 0.88 **<br>[0.81-0.97] | 0.90 **<br>[0.82-0.97] | 0.84 ****<br>[0.78-0.92] | 0.86 **<br>[0.78-0.94] | 0.93 ****<br>[0.89-0.96] |
| 41-50 | 1.31 **<br>[1.10-1.57] | 1.82 ****<br>[1.53-2.17] | 1.95 ****<br>[1.72-2.21] | 1.50 ****<br>[1.35-1.66] | 1.54 ****<br>[1.39-1.72] | 1.37 ****<br>[1.27-1.48] | 1.24 ****<br>[1.15-1.34] | 1.31 ****<br>[1.21-1.41] | 1.38 ****<br>[1.27-1.50] | 1.36 ****<br>[1.31-1.40] |
| 51-64 | 1.33 ****<br>[1.16-1.54] | 1.21 *<br>[1.03-1.42] | 1.46 ****<br>[1.31-1.63] | 1.39 ****<br>[1.28-1.52] | 1.50 ****<br>[1.38-1.64] | 1.36 ****<br>[1.28-1.44] | 1.33 ****<br>[1.26-1.41] | 1.34 ****<br>[1.26-1.41] | 1.39 ****<br>[1.31-1.48] | 1.35 ****<br>[1.32-1.39] |
| **Gender <sup>b</sup>** |  |  |  |  |  |  |  |  |  |  |
| Male | 1.12 *<br>[1.00-1.24] | 1.09<br>[0.97-1.22] | 1.08<br>[1.00-1.17] | 0.99<br>[0.93-1.06] | 0.97<br>[0.91-1.04] | 0.97<br>[0.92-1.02] | 0.98<br>[0.93-1.02] | 0.95 *<br>[0.91-0.99] | 0.92 **<br>[0.88-0.97] | 0.98 *<br>[0.96-1.00] |
| **Race <sup>c</sup>** |  |  |  |  |  |  |  |  |  |  |
| African American | 0.66 ****<br>[0.55-0.79] | 0.61 ****<br>[0.50-0.75] | 0.62 ****<br>[0.56-0.70] | 0.64 ****<br>[0.58-0.70] | 0.58 ****<br>[0.52-0.65] | 0.69 ****<br>[0.64-0.75] | 0.73 ****<br>[0.68-0.78] | 0.70 ****<br>[0.65-0.74] | 0.78 ****<br>[0.73-0.84] | 0.68 ****<br>[0.66-0.70] |
| Asian | 0.38 *<br>[0.17-0.85] | 0.15 **<br>[0.04-0.61] | 0.39 ***<br>[0.24-0.63] | 0.37 ****<br>[0.26-0.53] | 0.44 ****<br>[0.31-0.60] | 0.41 ****<br>[0.32-0.52] | 0.52 ****<br>[0.42-0.64] | 0.49 ****<br>[0.40-0.60] | 0.50 ****<br>[0.39-0.63] | 0.47 ****<br>[0.42-0.51] |
| Hispanic | 0.74<br>[0.49-1.12] | 0.57 *<br>[0.35-0.94] | 0.54 ***<br>[0.39-0.74] | 0.58 ***<br>[0.42-0.79] | 0.54 **<br>[0.34-0.84] | 0.74 *<br>[0.55-1.00] | 0.56 **<br>[0.39-0.81] | 0.68<br>[0.46-1.01] | 0.54 *<br>[0.33-0.87] | 0.53 ****<br>[0.46-0.60] |
| Native American | 0.65<br>[0.21-2.01] | 1.58<br>[0.65-3.83] | 0.56 *<br>[0.33-0.95] | 0.47 ***<br>[0.32-0.70] | 0.67 **<br>[0.50-0.88] | 0.79 *<br>[0.66-0.95] | 0.85<br>[0.71-1.01] | 1.17<br>[0.98-1.39] | 0.76 *<br>[0.61-0.94] | 0.84 ****<br>[0.77-0.91] |
| Others | 0.57 ****<br>[0.44-0.74] | 0.51 ****<br>[0.37-0.69] | 0.54 ****<br>[0.44-0.65] | 0.46 ****<br>[0.40-0.53] | 0.56 ****<br>[0.49-0.65] | 0.57 ****<br>[0.51-0.63] | 0.56 ****<br>[0.51-0.61] | 0.57 ****<br>[0.52-0.62] | 0.56 ****<br>[0.51-0.62] | 0.57 ****<br>[0.55-0.60] |
| **Marital Status <sup>d</sup>** |  |  |  |  |  |  |  |  |  |  |
| Divorced | 1.23 *<br>[1.00-1.50] | 1.19<br>[0.95-1.49] | 1.26 ***<br>[1.11-1.44] | 1.15 *<br>[1.03-1.28] | 1.14 *<br>[1.02-1.27] | 1.16 ***<br>[1.07-1.25] | 1.28 ****<br>[1.19-1.38] | 1.25 ****<br>[1.16-1.35] | 1.22 ****<br>[1.12-1.32] | 1.19 ****<br>[1.15-1.23] |
| Married | 0.65 ****<br>[0.56-0.75] | 0.72 ****<br>[0.61-0.85] | 0.74 ****<br>[0.67-0.82] | 0.71 ****<br>[0.65-0.77] | 0.69 ****<br>[0.64-0.75] | 0.70 ****<br>[0.66-0.74] | 0.75 ****<br>[0.71-0.80] | 0.82 ****<br>[0.77-0.86] | 0.82 ****<br>[0.77-0.87] | 0.73 ****<br>[0.71-0.75] |
| Widowed | 0.82<br>[0.66-1.03] | 0.66 **<br>[0.50-0.86] | 1.12<br>[0.96-1.29] | 0.90<br>[0.80-1.01] | 0.97<br>[0.86-1.09] | 0.94<br>[0.86-1.02] | 0.96<br>[0.89-1.05] | 0.96<br>[0.88-1.04] | 0.85 ***<br>[0.77-0.93] | 0.89 ****<br>[0.86-0.93] |
| Others | 0.97<br>[0.83-1.14] | 1.20 *<br>[1.02-1.42] | 0.79 **<br>[0.66-0.94] | 1.03<br>[0.89-1.20] | 0.93<br>[0.78-1.11] | 1.10<br>[0.99-1.22] | 1.12 *<br>[1.01-1.24] | 1.20 ***<br>[1.08-1.34] | 1.20 **<br>[1.07-1.35] | 0.92 ****<br>[0.88-0.96] |
| **Census Region <sup>e</sup>** |  |  |  |  |  |  |  |  |  |  |
| Midwest | 1.78 ****<br>[1.49-2.13] | 1.60 ****<br>[1.32-1.94] | 0.92<br>[0.83-1.02] | 1.11 *<br>[1.01-1.23] | 1.14 **<br>[1.03-1.26] | 1.15 ****<br>[1.08-1.24] | 0.97<br>[0.91-1.03] | 1.19 ****<br>[1.13-1.26] | 0.94<br>[0.88-1.01] | 1.01<br>[0.98-1.04] |
| Northeast | 2.39 ****<br>[2.03-2.80] | 2.12 ****<br>[1.78-2.52] | 1.17 **<br>[1.06-1.29] | 1.28 ****<br>[1.18-1.40] | 1.49 ****<br>[1.35-1.63] | 1.30 ****<br>[1.22-1.39] | 1.02<br>[0.96-1.09] | 0.84 ****<br>[0.79-0.90] | 0.96<br>[0.89-1.03] | 1.00<br>[0.97-1.02] |
| West | 2.59 ****<br>[2.04-3.31] | 1.47 *<br>[1.06-2.05] | 1.33 ****<br>[1.16-1.52] | 1.52 ****<br>[1.38-1.67] | 1.78 ****<br>[1.63-1.94] | 1.55 ****<br>[1.45-1.65] | 1.23 ****<br>[1.16-1.31] | 1.03<br>[0.97-1.09] | 1.13 **<br>[1.05-1.21] | 1.32 ****<br>[1.28-1.36] |
| **Urbanicity <sup>f</sup>** |  |  |  |  |  |  |  |  |  |  |
| Rural | 0.46 ****<br>[0.38-0.56] | 0.51 ****<br>[0.41-0.62] | 0.97<br>[0.87-1.08] | 0.91 *<br>[0.83-0.98] | 0.69 ****<br>[0.62-0.76] | 0.90 ***<br>[0.85-0.96] | 0.86 ****<br>[0.81-0.92] | 0.83 ****<br>[0.78-0.89] | 0.81 ****<br>[0.75-0.88] | 0.84 ****<br>[0.82-0.86] |

**a:** Reference - patients in 65+ age group. **b:** Reference - female patients. **c:** Reference - Caucasian patients. **d:** Reference - patients in single marital status. **e:** Reference - patients from South census region. **f:** Reference - patients in urban area. \* p < 0.05; \*\* p < 0.01; \*\*\* p < 0.001; \*\*\*\* p < 0.0001.

### 3.4 Prescription Opioid Exposures as Risk Factors for Opioid Overdose

Prescription opioids included for evaluation were listed in Table 3, along with their relevant properties and the overall prevalence between 2009 and 2017. Prescription opioid exposures as risk factors for opioid overdose were detailed in Table 4. Among MOR agonists, morphine (342.1‰) was most frequently prescribed, followed by fentanyl (297.5‰), oxycodone (263.3‰), hydromorphone (242.1‰) and hydrocodone (182.4‰), the high usage rates of which manifested the prescription opioid abuse [40]. The MOR antagonist, naloxone (157.8‰), was also commonly prescribed and compared to the non-exposed patients, patients exposed to naloxone (overall aOR: 3.24, 95% CI [3.18-3.31]; p < 0.0001) had a higher odds of opioid overdose, presumably due to its therapeutic effect to reverse overdose [3].

**Table 3.** Relevant Properties of Prescription Opioids and the Overall Prevalence (‰) between 2009 and 2017.

| **Prescription Opioids <sup>a</sup>** | **Physicochemical Properties <sup>b</sup>** |  |  | **Pharmacokinetic Properties <sup>c</sup>** |  |  | **Pharmacodynamic Properties <sup>d</sup>** |  |  | **Overall Prevalence** |
| --- | --- | --- | --- | --- | --- | --- | --- | --- | --- | --- |
|  | **logP** | **PSA** | **MW** | **MDR1** | **CYP2D6** | **CYP3A4** | **DOR** | **KOR** | **NMDAR** |  |
| **Agonist** |  |  |  |  |  |  |  |  |  |  |
| Morphine | 1.0 | 52.9 | 285.3 | + | - | + | + | + | - | 342.1 |
| Fentanyl | 4.1 | 23.6 | 336.5 | + | - | + | + | + | - | 297.5 |
| Oxycodone | 1.0 | 59.0 | 315.4 | - | + | + | + | + | - | 263.3 |
| Hydromorphone | 1.7 | 49.8 | 285.3 | - | - | - | + | + | - | 242.1 |
| Hydrocodone | 2.1 | 38.8 | 299.4 | - | + | + | + | - | - | 182.4 |
| Meperidine | 2.9 | 29.5 | 247.3 | - | + | + | - | + | + | 82.3 |
| Tramadol | 2.7 | 32.7 | 263.4 | + | + | + | + | + | + | 55.7 |
| Codeine | 1.2 | 41.9 | 299.4 | - | + | + | + | + | - | 43.9 |
| Loperamide | 4.4 | 43.8 | 477.0 | + | + | + | + | + | - | 23.1 |
| Diphenoxylate | 5.7 | 53.3 | 452.6 | - | - | - | + | - | - | 8.4 |
| Methadone | 4.1 | 20.3 | 309.4 | + | + | + | + | - | + | 7.8 |
| **Partial Agonist** |  |  |  |  |  |  |  |  |  |  |
| Nalbuphine | 2.0 | 73.2 | 357.4 | - | - | - | + | + | - | 45.7 |
| Butorphanol | 3.7 | 43.7 | 327.5 | - | - | - | + | + | - | 29.5 |
| Buprenorphine | 4.5 | 62.2 | 467.6 | + | + | + | + | + | - | 3.2 |
| **Antagonist** |  |  |  |  |  |  |  |  |  |  |
| Naloxone | 1.5 | 70.0 | 327.4 | - | - | + | + | + | - | 157.8 |

**a:** Prescription opioids are categorized by their action on opioid mu receptor (MOR), ordered by their overall prevalence. **b:** Relevant physicochemical properties include lipophilicity (logP), polar surface area (PSA) and molecular weight (MW). **c:** Relevant pharmacokinetic properties focus on whether the opioid interacts with 1) drug efflux transporter, multi-drug resistance protein 1 (MDR1) 2) metabolizing enzymes, cytochromes P450 2D6 (CYP2D6) and 3A4 (CYP3A4). **d:** Relevant pharmacodynamic properties focus on whether the opioid interacts with 1) other opioid receptors, delta opioid receptor (DOR) and kappa opioid receptor (KOR) 2) non-opioid receptor in modulating opioid analgesia, N-methyl-D-aspartate receptor (NMDAR). N.A. = Not Applicable.

**Table 4.** Prescription Opioid Exposures as Risk Factors for Opioid Overdose and Adjusted Odds Ratios from 2009 to 2017.

| Year | Adjusted Odds Ratios [95% CI] |  |  |  |  |  |  |  |  |  |
| --- | --- | --- | --- | --- | --- | --- | --- | --- | --- | --- |
|  | 2009 | 2010 | 2011 | 2012 | 2013 | 2014 | 2015 | 2016 | 2017 | Overall |
| **Agonist** |  |  |  |  |  |  |  |  |  |  |
| Morphine | 1.58 ****<br>[1.41-1.76] | 1.31 ****<br>[1.15-1.48] | 1.36 ****<br>[1.26-1.47] | 1.36 ****<br>[1.28-1.45] | 1.33 ****<br>[1.25-1.42] | 1.49 ****<br>[1.42-1.56] | 1.56 ****<br>[1.49-1.63] | 1.53 ****<br>[1.46-1.60] | 1.59 ****<br>[1.51-1.67] | 1.44 ****<br>[1.42-1.47] |
| Fentanyl | 1.05<br>[0.94-1.17] | 1.22 **<br>[1.07-1.38] | 1.23 ****<br>[1.14-1.33] | 1.24 ****<br>[1.16-1.32] | 1.56 ****<br>[1.45-1.68] | 1.27 ****<br>[1.22-1.33] | 1.33 ****<br>[1.27-1.39] | 1.47 ****<br>[1.40-1.54] | 1.51 ****<br>[1.42-1.60] | 1.22 ****<br>[1.20-1.25] |
| Oxycodone | 1.72 ****<br>[1.54-1.92] | 1.68 ****<br>[1.49-1.90] | 1.74 ****<br>[1.60-1.88] | 1.60 ****<br>[1.50-1.71] | 1.78 ****<br>[1.66-1.91] | 1.73 ****<br>[1.65-1.82] | 1.82 ****<br>[1.74-1.91] | 1.89 ****<br>[1.80-1.98] | 1.90 ****<br>[1.80-2.01] | 1.70 ****<br>[1.67-1.73] |
| Hydromorphone | 1.97 ****<br>[1.74-2.23] | 1.66 ****<br>[1.45-1.91] | 1.88 ****<br>[1.73-2.04] | 1.64 ****<br>[1.54-1.75] | 1.71 ****<br>[1.59-1.83] | 1.76 ****<br>[1.68-1.84] | 1.79 ****<br>[1.71-1.87] | 1.95 ****<br>[1.86-2.04] | 1.89 ****<br>[1.80-1.99] | 1.81 ****<br>[1.77-1.84] |
| Hydrocodone | 1.24 **<br>[1.08-1.42] | 1.01 <sup>a</sup><br>[0.85-1.20] | 1.19 ***<br>[1.08-1.30] | 1.31 ****<br>[1.22-1.41] | 1.17 ****<br>[1.08-1.27] | 1.26 ****<br>[1.20-1.33] | 1.23 ****<br>[1.17-1.30] | 1.25 ****<br>[1.18-1.33] | 1.18 ****<br>[1.10-1.27] | 1.12 ****<br>[1.09-1.15] |
| Meperidine | 1.23<br>[0.94-1.61] | 0.98 <sup>a</sup><br>[0.64-1.50] | 1.09<br>[0.96-1.24] | 1.13 *<br>[1.02-1.30] | 1.15 *<br>[1.02-1.30] | 1.07<br>[0.99-1.15] | 1.08 *<br>[1.00-1.16] | 1.14 **<br>[1.05-1.23] | 1.01 <sup>a</sup><br>[0.93-1.11] | 1.09 ****<br>[1.06-1.13] |
| Tramadol | 3.70 ****<br>[3.10-4.41] | 3.47 ****<br>[2.86-4.20] | 3.01 ****<br>[2.68-3.38] | 2.69 ****<br>[2.45-2.95] | 2.11 ****<br>[1.91-2.34] | 2.21 ****<br>[2.08-2.36] | 2.16 ****<br>[2.03-2.30] | 1.88 ****<br>[1.76-2.01] | 1.50 ****<br>[1.36-1.65] | 2.20 ****<br>[2.14-2.27] |
| Codeine | 0.96<br>[0.75-1.22] | 0.81<br>[0.58-1.13] | 0.96<br>[0.79-1.15] | 0.95<br>[0.81-1.12] | 0.71 ***<br>[0.58-0.85] | 0.89<br>[0.78-1.01] | 0.89 *<br>[0.78-1.00] | 0.69 ****<br>[0.60-0.80] | 0.73 **<br>[0.60-0.89] | 0.78 ****<br>[0.74-0.83] |
| Loperamide | 2.08 ****<br>[1.57-2.75] | 1.41<br>[0.95-2.10] | 2.20 ****<br>[1.85-2.61] | 2.16 ****<br>[1.87-2.49] | 2.11 ****<br>[1.80-2.47] | 2.12 ****<br>[1.92-2.35] | 2.17 ****<br>[1.98-2.39] | 1.96 ****<br>[1.77-2.17] | 1.63 ****<br>[1.44-1.85] | 2.03 ****<br>[1.94-2.12] |
| Diphenoxylate | 1.63 *<br>[1.01-2.64] | 1.69<br>[0.98-2.93] | 2.06 ****<br>[1.56-2.72] | 1.88 ****<br>[1.45-2.43] | 1.57 **<br>[1.16-2.12] | 1.33 **<br>[1.10-1.62] | 1.39 ***<br>[1.16-1.67] | 1.69 ****<br>[1.37-2.09] | 1.79 ****<br>[1.38-2.32] | 1.48 ****<br>[1.36-1.61] |
| Methadone | 7.86 ****<br>[6.34-9.74] | 6.36 ****<br>[4.88-8.28] | 7.12 ****<br>[6.10-8.32] | 7.07 ****<br>[6.23-8.03] | 6.59 ****<br>[5.66-7.67] | 6.78 ****<br>[6.14-7.49] | 6.47 ****<br>[5.86-7.1] | 6.53 ****<br>[5.85-7.30] | 5.46 ****<br>[4.78-6.22] | 6.51 ****<br>[6.23-6.80] |
| **Partial Agonist** |  |  |  |  |  |  |  |  |  |  |
| Nalbuphine | 0.38 ****<br>[0.26-0.57] | 0.38 ****<br>[0.25-0.60] | 0.46 ****<br>[0.34-0.61] | 0.54 ****<br>[0.43-0.66] | 0.63 ****<br>[0.50-0.79] | 0.46 ****<br>[0.39-0.54] | 0.57 ****<br>[0.50-0.66] | 0.52 ****<br>[0.45-0.60] | 0.91 ****<br>[0.78-1.07] | 0.56 ****<br>[0.53-0.60] |
| Butorphanol | 0.19 ****<br>[0.09-0.44] | 0.08 ***<br>[0.02-0.34] | 0.10 ****<br>[0.05-0.19] | 0.13 ****<br>[0.06-0.29] | 0.06 ****<br>[0.03-0.14] | 0.11 ****<br>[0.08-0.17] | 0.05 ****<br>[0.03-0.08] | 0.11 ****<br>[0.06-0.18] | 0.08 ****<br>[0.04-0.16] | 0.09 ****<br>[0.07-0.11] |
| Buprenorphine | 3.14 ****<br>[1.80-5.46] | 1.90<br>[0.78-4.61] | 2.33 ****<br>[1.58-3.44] | 3.37 ****<br>[2.48-4.60] | 5.14 ****<br>[3.73-7.10] | 3.35 ****<br>[2.73-4.11] | 2.77 ****<br>[2.19-3.49] | 2.56 ****<br>[2.05-3.21] | 1.74 ****<br>[1.29-2.35] | 2.89 ****<br>[2.63-3.19] |
| **Antagonist** |  |  |  |  |  |  |  |  |  |  |
| Naloxone | 6.40 ****<br>[5.70-7.19] | 5.70 ****<br>[5.00-6.51] | 4.19 ****<br>[3.85-4.55] | 3.93 ****<br>[3.68-4.20] | 2.80 ****<br>[2.61-3.01] | 3.50 ****<br>[3.34-3.67] | 3.13 ****<br>[3.00-3.28] | 2.49 ****<br>[2.38-2.61] | 2.53 ****<br>[2.39-2.67] | 3.24 ****<br>[3.18-3.31] |

a: Power less than 80%. \* p < 0.05; \*\* p < 0.01; \*\*\* p < 0.001; \*\*\*\* p < 0.0001. N.A. = Not Applicable.

For MOR partial agonists, buprenorphine (3.2‰) was less frequently prescribed than nalbuphine (45.7‰) and butorphanol (29.5‰). Besides, exposures to partial agonists, nalbuphine (overall aOR: 0.565, 95% CI [0.53-0.60]; p < 0.0001) and butorphanol (aOR: 0.09, 95% CI [0.07 - 0.11]; p < 0.0001), were associated with reduced odds of overdose whereas buprenorphine (overall aOR: 2.89, 95% CI [2.63-3.19]; p < 0.0001) was positively associated with opioid overdose. This could be explained by that buprenorphine is for medication assisted treatment (MAT), which is closely related to opioid overdose [41]. Exposures to opioid agonists were also positively associated with opioid overdose except for codeine (overall aOR: 0.78, 95% CI [0.74-0.83]; p < 0.0001), consistent with a previous finding that codeine had lowest risk for severe respiratory depression [13]. Moreover, different opioid agonists posed unique aORs, reflecting varied association strengths in opioid overdose.

### 3.5 Optimal Properties for Better Opioid Agonists

To explore the optimal properties underlying better opioid agonists with less overdose effects, we examined the relationships between their relevant properties (Table 3) and the yearly aORs of opioid agonists (Table 4), depicted in Fig. 1. Note that opioid agonists targeting for peripheral MORs, namely loperamide and diphenoxylate, and methadone for MAT purpose, were excluded [41, 42]. For the physicochemical properties, lower aORs, i.e., weakened association strengths, were observed for opioid agonists with high MW (median decreased from 1.62 to 1.24; p < 0.001). The interpretation is that when an opioid agonist’s MW is high, its association with opioid overdose would be weakened, possibly due to reduced BBB permeability [33]. For logP (median decreased from 1.49 to 1.40; p = 0.2) and PSA (median increased from 1.25 to 1.60; p = 0.45), no significant associations were observed. From the pharmacokinetic perspective, a negative interaction with MDR1 was associated with lower aORs (median decreased from 1.51 to 1.25; p < 0.05), which indicates that if an opioid agonist does not interact with MDR1, its association strength with opioid overdose would be lowered. This could be due to that an interaction between opioids and MDR1 at the BBB can up-regulate the expression of MDR1, which leads to opioid tolerance, abuse, dependence and eventually overdose [43, 44]. For the interaction with CYP3A4, a positive interaction was associated with lower aORs (median decreased from 1.79 to 1.32; p < 0.01), which suggests that CYP3A4 metabolism may be protective for opioid agonists from overdose as a result of reduced bioavailability during first-pass metabolism [35]. From the pharmacodynamic perspective, lower aORs were observed when opioid agonists do not interact with DOR (median decreased from 1.52 to 1.13; p < 0.01) or KOR (median decreased from 1.53 to 1.23; p = 0.07). The interpretation is that when opioid agonists are selective towards MOR, overdose events would be reduced. For NMDAR (median increased from 1.36 to 1.69; p = 0.16), no significant difference in aORs was observed.

**Fig. 1.**
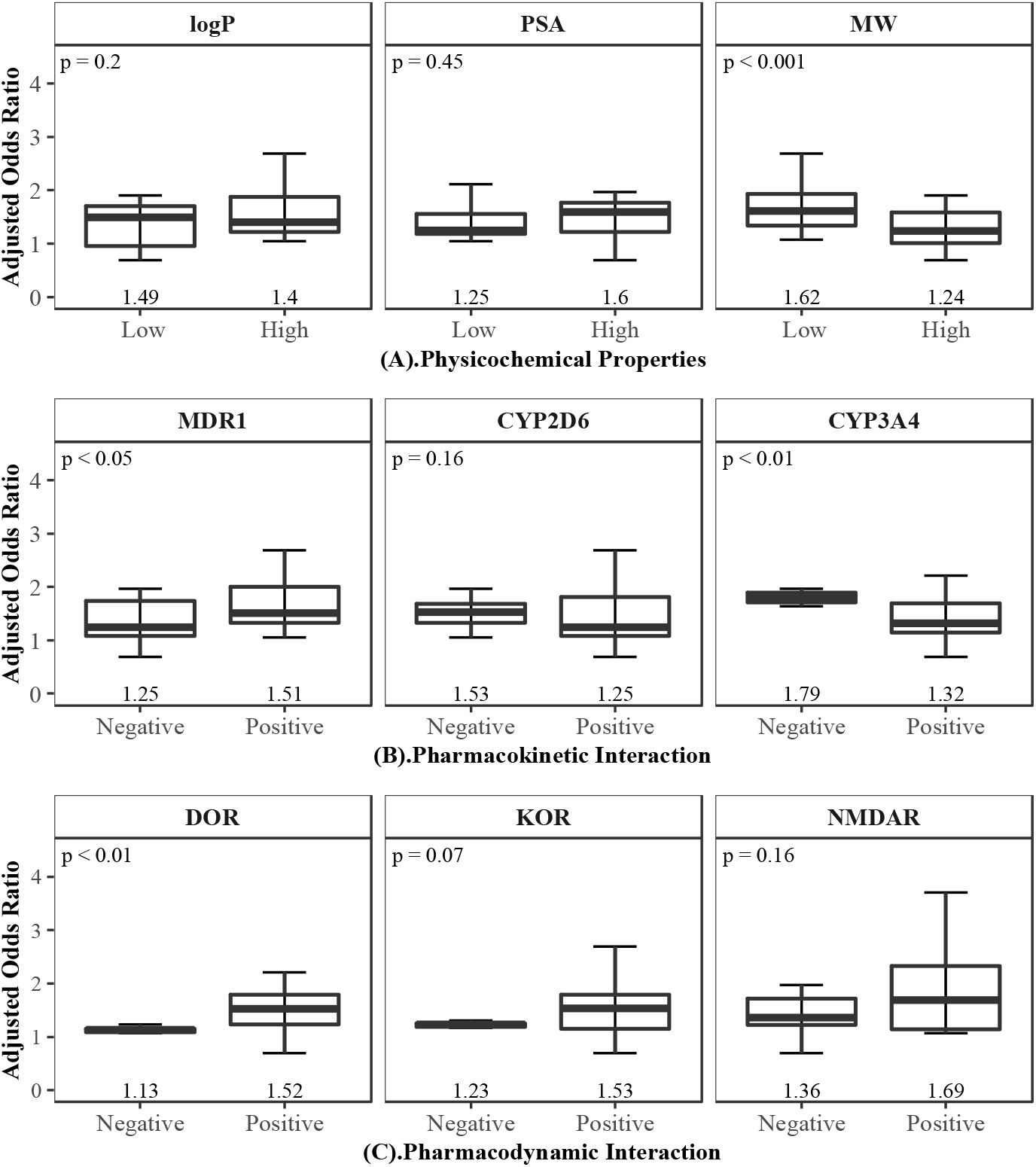
Relationships between Innate Properties of Opioid Agonists and the Adjusted Odds Ratios in Opioid Overdose. **(A)**. Wilcoxon test on opioid agonists’ aORs in opioid overdose and their discretized physicochemical properties: lipophilicity (logP), polar surface area (PSA) and molecular weight (MW). **(B)**. Wilcoxon test on opioid agonists’ aORs in opioid overdose and their interaction type with efflux transporter and metabolizing enzymes: multi-drug resistance protein 1 (MDR1), cytochromes P450 2D6 (CYP2D6) and 3A4 (CYP3A4). (**C)**. Wilcoxon test on opioid agonists’ aORs in opioid overdose and their interaction type with receptors: delta opioid receptor (DOR), kappa opioid receptor (KOR) and N-methyl-D-aspartate receptor (NMDAR).

## 4 Discussion

### 4.1 An Ongoing Need for Targeted Overdose Prevention

In this study, we found that opioid overdose increased significantly at the US inpatient care setting from 2009 to 2017. As pointed out by Danovitch et al [45], inpatient opioid overdose is a serious harm, yet preventable, and is likely to be underestimated in much of current literature. The significant increasing trends observed from Health Facts^®^, a large-scale EHRs database, indicated that prevention and control measures for opioid overdose had been inadequate at the US inpatient setting over these years, especially for patient subgroups with prominent fold increases, such as patients from South and patients in rural area.

We also quantified how patient demographics were associated with inpatient opioid overdose and found that certain demographics increased the odds of opioid overdose. For instance, compared to the major demographic counterpart age 65+, patients aged 51-64 and 41-50 were more likely to have opioid overdose, which was partly consistent with a recent Mortality Disparities in American Communities study, where patients aged 40-59 were attributed to the most part of opioid overdose deaths from 2008 to 2015 [46]. Patients in urban area had higher odds of opioid overdose than those in rural area, which reflected the urbanicity of the opioid overdose epidemic [46]. The associated risk factors suggest the need for targeted measures in patient subgroups at higher risk.

### 4.2 A Necessity to Differentiate Prescription Opioids

In addition to patient demographics, prescription opioid exposures were also evaluated as risk factors. Our study showed that prescription opioids were commonly prescribed. For instance, 342.1‰ patients were prescribed with medications containing morphine, the high prevalence of which manifested inappropriate opioid prescribing practice and, eventually, prevalent opioid overdose [47]. Previous studies showed that opioid overdose was associated with opioid prescription patterns, such as dosage and duration [48-51]. Yet, measures to reduce prescription opioids supply alone may not be enough to combat the opioid crisis [52].

In current literature, prescription opioids were usually aggregated for the morphine milligram equivalents (MME) calculation, a widely adopted metric [16-18]. One potential misconception arising from MME is that all opioids are interchangeable, which can falsely justify the transitioning of one opioid to other “equivalent” opioids [53, 54]. A previous study based on a cohort of 307 patients showed that some prescription opioids were associated with a much higher risk for severe respiratory depression than others [13]. For example, fentanyl had 20-fold higher relative risk compared to codeine, the lowest-risk opioid. In fact, a wide range of differences exist among prescription opioids. For instance, during distribution to the site of action, i.e., MOR in the central nervous system, they exhibit varied BBB permeability [55]. Besides, opioids also have disparate binding profiles to the opioid receptors [56, 57]. Our study revealed varied associations with opioid overdose for different prescription opioids, indicating a necessity to differentiate prescription opioids in opioid prescribing practice [13].

### 4.2 Real-World Evidence on Optimal Properties for Better Opioids

Given the foreseeable continuing trends of opioid overdose in the US and the high prevalence of prescription opioids for pain, there is an undisputed need to develop better opioids with less overdose effects. Our study generated RWEs on optimal properties for better opioid agonists. For instance, from the physicochemical perspective, opioid agonists with low MW had significantly higher association strengths with opioid overdose, possibly due to increased BBB permeability. From pharmacokinetic and pharmacodynamic perspectives, when opioid agonists interact with MDR1, DOR or KOR, the association strengths in opioid overdose were significantly increased. Thus, the interactions with the efflux transporter and other opioid receptors should be taken into consideration during early drug discovery for better opioid agonists. Although some optimal properties have been proposed before, they were mostly based on preclinical studies and there still lacks a clear link to the safety profiles in human. With the emergence of artificial intelligence in drug discovery, molecule structure design biased towards pre-defined attributes has become a reality [58, 59]. The RWEs on the optimal properties for opioids with less overdose effects can function as actionable knowledge to develop better opioids [26].

### 4.3 Limitations

Our study also had some limitations. Firstly, we only used ICD-9 /ICD-10 codes to identify patients with opioid overdose, which may not be fully accurate [60, 61]. Besides, there was a transition of ICD-9 codes to ICD-10 codes in 2015, which can lead to discontinuities in the opioid overdose trends [62]. Secondly, we only focused on patient demographics and prescription opioid exposures as risk factors for opioid overdose. Other potential risk factors like comorbidities and surgery procedures were not examined yet [63, 64]. In addition, opioid prescription patterns including dosage and duration also need further assessment. Thirdly, because of the observational study design, where we did not strictly distinguish the temporal order of being prescribed with a medication and getting diagnosed as opioid overdose, the association strengths, thus, may not necessarily represent causality. Lastly, our study was based on a single database. Exploitation on other RWD sources would also be needed.

Nevertheless, despite the limitations, we conducted a comprehensive study on opioid overdose through investigating its patterns and generating RWEs on the optimal properties for better opioids, which provided critical knowledge to combat the opioid overdose epidemic.

## 5 Conclusions

A retrospective observational study was conducted using a large-scale EHRs database to examine the opioid overdose patterns, including temporal trends and associated risk factors. The significant increasing trends of opioid overdose from 2009 to 2017 indicated that prevention and control measures for opioid overdose had been inadequate at the US inpatient care setting. Besides, patient demographics and prescription opioid exposures were associated with higher odds of opioid overdose, suggesting that targeted prevention and control measures are needed. Furthermore, there are optimal properties underlying opioid agonists with reduced overdose effects, which hold great potential for the development of better opioids.

## Data Availability

The data used for analysis is from Cerner Health Facts.

https://www.cerner.com/

## Notes

### Competing Interest Statement

The authors have declared no competing interest.

### Funding Statement

This work was funded partially by the Stony Brook University OVPR Seed Grant 1158484-63845-6.

### Author Declarations

Health Facts is a de-identified database. The use of the Health Facts data for research purposes is approved by Cerner and Stony Brook Medicine.

### Summary of Updates

Major revisions include the following. 1. The abstract is revised. 2. Three key points are added.

