## Supporting Information for "A Large-Scale Observational Study on the Temporal Trends and Risk Factors of Opioid Overdose: Real-World Evidence for Better Opioids" (from another version of this preprint: version 1, 2020-10-11)

### Table S1. Diagnosis Codes for Opioid Overdose from International Classification of Diseases (ICD) Ninth and Tenth Revision

| **Version ^a^** | **Condition** | **Code** | **Code Description** |
| --- | --- | --- | --- |
| ICD-9 | opioid poisoning | 965.0 | Poisoning by opiates and related narcotics |
| ICD-9 | opioid poisoning | 965.00 | Poisoning by opium (alkaloids), unspecified |
| ICD-9 | opioid poisoning | 965.01 | Poisoning by heroin |
| ICD-9 | opioid poisoning | 965.02 | Poisoning by methadone |
| ICD-9 | opioid poisoning | 965.09 | Poisoning by other opiates or related narcotics |
| ICD-9 | opioid poisoning | 970.1 | Poisoning by opiate antagonists |
| ICD-9 | opioid poisoning | E850.0 | Accidental poisoning by heroin |
| ICD-9 | opioid poisoning | E850.1 | Accidental poisoning by methadone |
| ICD-9 | opioid poisoning | E850.2 | Accidental poisoning by other opiates or related narcotics |
| ICD-9 | opioid adverse effect | E935.0 | Heroin causing adverse effects in therapeutic use |
| ICD-9 | opioid adverse effect | E935.1 | Methadone causing adverse effects in therapeutic use |
| ICD-9 | opioid adverse effect | E935.2 | Other opiates and related narcotics causing adverse effects in therapeutic use |
| ICD-9 | opioid adverse effect | E940.1 | Opiate antagonists causing adverse effects in therapeutic use |
| ICD-10 | opioid poisoning | T40.0X1 | Poisoning by opium, accidental (unintentional) |
| ICD-10 | opioid poisoning | T40.0X1A | Poisoning by opium, accidental (unintentional), initial encounter |
| ICD-10 | opioid poisoning | T40.0X1D | Poisoning by opium, accidental (unintentional), subsequent encounter |
| ICD-10 | opioid poisoning | T40.0X1S | Poisoning by opium, accidental (unintentional), sequela |
| ICD-10 | opioid poisoning | T40.0X2 | Poisoning by opium, intentional self-harm |
| ICD-10 | opioid poisoning | T40.0X2A | Poisoning by opium, intentional self-harm, initial encounter |
| ICD-10 | opioid poisoning | T40.0X2D | Poisoning by opium, intentional self-harm, subsequent encounter |
| ICD-10 | opioid poisoning | T40.0X2S | Poisoning by opium, intentional self-harm, sequela |
| ICD-10 | opioid poisoning | T40.0X3 | Poisoning by opium, assault |
| ICD-10 | opioid poisoning | T40.0X3A | Poisoning by opium, assault, initial encounter |
| ICD-10 | opioid poisoning | T40.0X3D | Poisoning by opium, assault, subsequent encounter |
| ICD-10 | opioid poisoning | T40.0X3S | Poisoning by opium, assault, sequela |
| ICD-10 | opioid poisoning | T40.0X4 | Poisoning by opium, undetermined |
| ICD-10 | opioid poisoning | T40.0X4A | Poisoning by opium, undetermined, initial encounter |
| ICD-10 | opioid poisoning | T40.0X4D | Poisoning by opium, undetermined, subsequent encounter |
| ICD-10 | opioid poisoning | T40.0X4S | Poisoning by opium, undetermined, sequela |
| ICD-10 | opioid adverse effect | T40.0X5 | Adverse effect of opium |
| ICD-10 | opioid adverse effect | T40.0X5A | Adverse effect of opium, initial encounter |
| ICD-10 | opioid adverse effect | T40.0X5D | Adverse effect of opium, subsequent encounter |
| ICD-10 | opioid adverse effect | T40.0X5S | Adverse effect of opium, sequela |
| ICD-10 | opioid poisoning | T40.1X1 | Poisoning by heroin, accidental (unintentional) |
| ICD-10 | opioid poisoning | T40.1X1A | Poisoning by heroin, accidental (unintentional), initial encounter |
| ICD-10 | opioid poisoning | T40.1X1D | Poisoning by heroin, accidental (unintentional), subsequent encounter |
| ICD-10 | opioid poisoning | T40.1X1S | Poisoning by heroin, accidental (unintentional), sequela |
| ICD-10 | opioid poisoning | T40.1X2 | Poisoning by heroin, intentional self-harm |
| ICD-10 | opioid poisoning | T40.1X2A | Poisoning by heroin, intentional self-harm, initial encounter |
| ICD-10 | opioid poisoning | T40.1X2D | Poisoning by heroin, intentional self-harm, subsequent encounter |
| ICD-10 | opioid poisoning | T40.1X2S | Poisoning by heroin, intentional self-harm, sequela |
| ICD-10 | opioid poisoning | T40.1X3 | Poisoning by heroin, assault |
| ICD-10 | opioid poisoning | T40.1X3A | Poisoning by heroin, assault, initial encounter |
| ICD-10 | opioid poisoning | T40.1X3D | Poisoning by heroin, assault, subsequent encounter |
| ICD-10 | opioid poisoning | T40.1X3S | Poisoning by heroin, assault, sequela |
| ICD-10 | opioid poisoning | T40.1X4 | Poisoning by heroin, undetermined |
| ICD-10 | opioid poisoning | T40.1X4A | Poisoning by heroin, undetermined, initial encounter |
| ICD-10 | opioid poisoning | T40.1X4D | Poisoning by heroin, undetermined, subsequent encounter |
| ICD-10 | opioid poisoning | T40.1X4S | Poisoning by heroin, undetermined, sequela |
| ICD-10 | opioid poisoning | T40.2X1 | Poisoning by other opioids, accidental (unintentional) |
| ICD-10 | opioid poisoning | T40.2X1A | Poisoning by other opioids, accidental (unintentional), initial encounter |
| ICD-10 | opioid poisoning | T40.2X1D | Poisoning by other opioids, accidental (unintentional), subsequent encounter |
| ICD-10 | opioid poisoning | T40.2X1S | Poisoning by other opioids, accidental (unintentional), sequela |
| ICD-10 | opioid poisoning | T40.2X2 | Poisoning by other opioids, intentional self-harm |
| ICD-10 | opioid poisoning | T40.2X2A | Poisoning by other opioids, intentional self-harm, initial encounter |
| ICD-10 | opioid poisoning | T40.2X2D | Poisoning by other opioids, intentional self-harm, subsequent encounter |
| ICD-10 | opioid poisoning | T40.2X2S | Poisoning by other opioids, intentional self-harm, sequela |
| ICD-10 | opioid poisoning | T40.2X3 | Poisoning by other opioids, assault |
| ICD-10 | opioid poisoning | T40.2X3A | Poisoning by other opioids, assault, initial encounter |
| ICD-10 | opioid poisoning | T40.2X3D | Poisoning by other opioids, assault, subsequent encounter |
| ICD-10 | opioid poisoning | T40.2X3S | Poisoning by other opioids, assault, sequela |
| ICD-10 | opioid poisoning | T40.2X4 | Poisoning by other opioids, undetermined |
| ICD-10 | opioid poisoning | T40.2X4A | Poisoning by other opioids, undetermined, initial encounter |
| ICD-10 | opioid poisoning | T40.2X4D | Poisoning by other opioids, undetermined, subsequent encounter |
| ICD-10 | opioid poisoning | T40.2X4S | Poisoning by other opioids, undetermined, sequela |
| ICD-10 | opioid adverse effect | T40.2X5 | Adverse effect of other opioids |
| ICD-10 | opioid adverse effect | T40.2X5A | Adverse effect of other opioids, initial encounter |
| ICD-10 | opioid adverse effect | T40.2X5D | Adverse effect of other opioids, subsequent encounter |
| ICD-10 | opioid adverse effect | T40.2X5S | Adverse effect of other opioids, sequela |
| ICD-10 | opioid poisoning | T40.3X1 | Poisoning by methadone, accidental (unintentional) |
| ICD-10 | opioid poisoning | T40.3X1A | Poisoning by methadone, accidental (unintentional), initial encounter |
| ICD-10 | opioid poisoning | T40.3X1D | Poisoning by methadone, accidental (unintentional), subsequent encounter |
| ICD-10 | opioid poisoning | T40.3X1S | Poisoning by methadone, accidental (unintentional), sequela |
| ICD-10 | opioid poisoning | T40.3X2 | Poisoning by methadone, intentional self-harm |
| ICD-10 | opioid poisoning | T40.3X2A | Poisoning by methadone, intentional self-harm, initial encounter |
| ICD-10 | opioid poisoning | T40.3X2D | Poisoning by methadone, intentional self-harm, subsequent encounter |
| ICD-10 | opioid poisoning | T40.3X2S | Poisoning by methadone, intentional self-harm, sequela |
| ICD-10 | opioid poisoning | T40.3X3 | Poisoning by methadone, assault |
| ICD-10 | opioid poisoning | T40.3X3A | Poisoning by methadone, assault, initial encounter |
| ICD-10 | opioid poisoning | T40.3X3D | Poisoning by methadone, assault, subsequent encounter |
| ICD-10 | opioid poisoning | T40.3X3S | Poisoning by methadone, assault, sequela |
| ICD-10 | opioid poisoning | T40.3X4 | Poisoning by methadone, undetermined |
| ICD-10 | opioid poisoning | T40.3X4A | Poisoning by methadone, undetermined, initial encounter |
| ICD-10 | opioid poisoning | T40.3X4D | Poisoning by methadone, undetermined, subsequent encounter |
| ICD-10 | opioid poisoning | T40.3X4S | Poisoning by methadone, undetermined, sequela |
| ICD-10 | opioid adverse effect | T40.3X5 | Adverse effect of methadone |
| ICD-10 | opioid adverse effect | T40.3X5A | Adverse effect of methadone, initial encounter |
| ICD-10 | opioid adverse effect | T40.3X5D | Adverse effect of methadone, subsequent encounter |
| ICD-10 | opioid adverse effect | T40.3X5S | Adverse effect of methadone, sequela |
| ICD-10 | opioid poisoning | T40.4X1 | Poisoning by other synthetic narcotics, accidental (unintentional) |
| ICD-10 | opioid poisoning | T40.4X1A | Poisoning by other synthetic narcotics, accidental (unintentional), initial encounter |
| ICD-10 | opioid poisoning | T40.4X1D | Poisoning by other synthetic narcotics, accidental (unintentional), subsequent encounter |
| ICD-10 | opioid poisoning | T40.4X1S | Poisoning by other synthetic narcotics, accidental (unintentional), sequela |
| ICD-10 | opioid poisoning | T40.4X2 | Poisoning by other synthetic narcotics, intentional self-harm |
| ICD-10 | opioid poisoning | T40.4X2A | Poisoning by other synthetic narcotics, intentional self-harm, initial encounter |
| ICD-10 | opioid poisoning | T40.4X2D | Poisoning by other synthetic narcotics, intentional self-harm, subsequent encounter |
| ICD-10 | opioid poisoning | T40.4X2S | Poisoning by other synthetic narcotics, intentional self-harm, sequela |
| ICD-10 | opioid poisoning | T40.4X3 | Poisoning by other synthetic narcotics, assault |
| ICD-10 | opioid poisoning | T40.4X3A | Poisoning by other synthetic narcotics, assault, initial encounter |
| ICD-10 | opioid poisoning | T40.4X3D | Poisoning by other synthetic narcotics, assault, subsequent encounter |
| ICD-10 | opioid poisoning | T40.4X3S | Poisoning by other synthetic narcotics, assault, sequela |
| ICD-10 | opioid poisoning | T40.4X4 | Poisoning by other synthetic narcotics, undetermined |
| ICD-10 | opioid poisoning | T40.4X4A | Poisoning by other synthetic narcotics, undetermined, initial encounter |
| ICD-10 | opioid poisoning | T40.4X4D | Poisoning by other synthetic narcotics, undetermined, subsequent encounter |
| ICD-10 | opioid poisoning | T40.4X4S | Poisoning by other synthetic narcotics, undetermined, sequela |
| ICD-10 | opioid adverse effect | T40.4X5 | Adverse effect of other synthetic narcotics |
| ICD-10 | opioid adverse effect | T40.4X5A | Adverse effect of other synthetic narcotics, initial encounter |
| ICD-10 | opioid adverse effect | T40.4X5D | Adverse effect of other synthetic narcotics, subsequent encounter |
| ICD-10 | opioid adverse effect | T40.4X5S | Adverse effect of other synthetic narcotics, sequela |
| ICD-10 | opioid poisoning | T40.601 | Poisoning by unspecified narcotics, accidental (unintentional) |
| ICD-10 | opioid poisoning | T40.601A | Poisoning by unspecified narcotics, accidental (unintentional), initial encounter |
| ICD-10 | opioid poisoning | T40.601D | Poisoning by unspecified narcotics, accidental (unintentional), subsequent encounter |
| ICD-10 | opioid poisoning | T40.601S | Poisoning by unspecified narcotics, accidental (unintentional), sequela |
| ICD-10 | opioid poisoning | T40.602 | Poisoning by unspecified narcotics, intentional self-harm |
| ICD-10 | opioid poisoning | T40.602A | Poisoning by unspecified narcotics, intentional self-harm, initial encounter |
| ICD-10 | opioid poisoning | T40.602D | Poisoning by unspecified narcotics, intentional self-harm, subsequent encounter |
| ICD-10 | opioid poisoning | T40.602S | Poisoning by unspecified narcotics, intentional self-harm, sequela |
| ICD-10 | opioid poisoning | T40.603 | Poisoning by unspecified narcotics, assault |
| ICD-10 | opioid poisoning | T40.603A | Poisoning by unspecified narcotics, assault, initial encounter |
| ICD-10 | opioid poisoning | T40.603D | Poisoning by unspecified narcotics, assault, subsequent encounter |
| ICD-10 | opioid poisoning | T40.603S | Poisoning by unspecified narcotics, assault, sequela |
| ICD-10 | opioid poisoning | T40.604 | Poisoning by unspecified narcotics, undetermined |
| ICD-10 | opioid poisoning | T40.604A | Poisoning by unspecified narcotics, undetermined, initial encounter |
| ICD-10 | opioid poisoning | T40.604D | Poisoning by unspecified narcotics, undetermined, subsequent encounter |
| ICD-10 | opioid poisoning | T40.604S | Poisoning by unspecified narcotics, undetermined, sequela |
| ICD-10 | opioid adverse effect | T40.605 | Adverse effect of unspecified narcotics |
| ICD-10 | opioid adverse effect | T40.605A | Adverse effect of unspecified narcotics, initial encounter |
| ICD-10 | opioid adverse effect | T40.605D | Adverse effect of unspecified narcotics, subsequent encounter |
| ICD-10 | opioid adverse effect | T40.605S | Adverse effect of unspecified narcotics, sequela |
| ICD-10 | opioid poisoning | T40.691 | Poisoning by other narcotics, accidental (unintentional) |
| ICD-10 | opioid poisoning | T40.691A | Poisoning by other narcotics, accidental (unintentional), initial encounter |
| ICD-10 | opioid poisoning | T40.691D | Poisoning by other narcotics, accidental (unintentional), subsequent encounter |
| ICD-10 | opioid poisoning | T40.691S | Poisoning by other narcotics, accidental (unintentional), sequela |
| ICD-10 | opioid poisoning | T40.692 | Poisoning by other narcotics, intentional self-harm |
| ICD-10 | opioid poisoning | T40.692A | Poisoning by other narcotics, intentional self-harm, initial encounter |
| ICD-10 | opioid poisoning | T40.692D | Poisoning by other narcotics, intentional self-harm, subsequent encounter |
| ICD-10 | opioid poisoning | T40.692S | Poisoning by other narcotics, intentional self-harm, sequela |
| ICD-10 | opioid poisoning | T40.693 | Poisoning by other narcotics, assault |
| ICD-10 | opioid poisoning | T40.693A | Poisoning by other narcotics, assault, initial encounter |
| ICD-10 | opioid poisoning | T40.693D | Poisoning by other narcotics, assault, subsequent encounter |
| ICD-10 | opioid poisoning | T40.693S | Poisoning by other narcotics, assault, sequela |
| ICD-10 | opioid poisoning | T40.694 | Poisoning by other narcotics, undetermined |
| ICD-10 | opioid poisoning | T40.694A | Poisoning by other narcotics, undetermined, initial encounter |
| ICD-10 | opioid poisoning | T40.694D | Poisoning by other narcotics, undetermined, subsequent encounter |
| ICD-10 | opioid poisoning | T40.694S | Poisoning by other narcotics, undetermined, sequela |
| ICD-10 | opioid adverse effect | T40.695 | Adverse effect of other narcotics |
| ICD-10 | opioid adverse effect | T40.695A | Adverse effect of other narcotics, initial encounter |
| ICD-10 | opioid adverse effect | T40.695D | Adverse effect of other narcotics, subsequent encounter |
| ICD-10 | opioid adverse effect | T40.695S | Adverse effect of other narcotics, sequela |

a: The transition from ICD-9 codes to ICD-10 codes took effect in October 2015 ^1^.

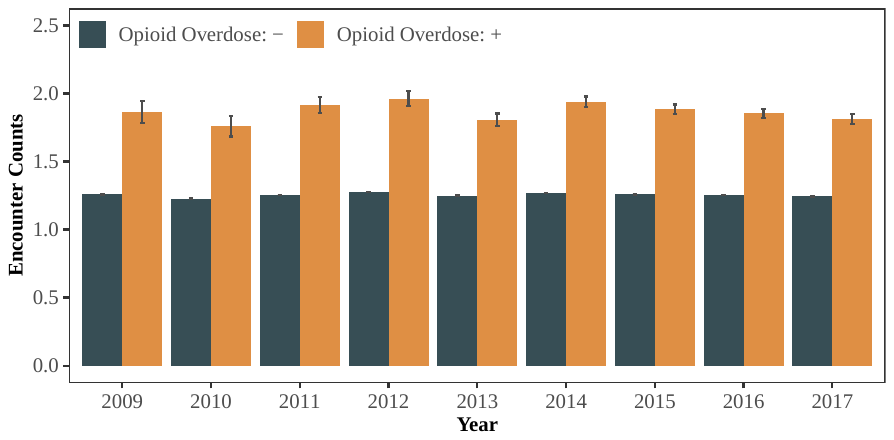

### Figure S1. Inpatient Encounter Counts for Patients from 2009 to 2017

**Notes:** Bar represents mean and error bar represents 95% confidence interval.
